# Figure accessibility for readers with colour vision deficiency: analysis of leading medical journals

**DOI:** 10.64898/2025.12.11.25342084

**Authors:** Kathryn Albany-Ward, Yaning Wu

## Abstract

Colour vision deficiency (CVD) affects up to 8% of males and 0.5% of females and currently lacks effective treatments. Individuals with CVD require visually accessible environments to flourish, the absence of which can cause unacceptable disparities in academic achievement and career progression. For current and aspiring clinician-scientists with CVD, inaccessible colour figures in scientific publications can hinder understanding of key information and potentially cause patient harm. Therefore, our work characterises the accessibility of major medical journal figures for individuals with CVD using established guidelines and provides recommendations for improving figure accessibility.

We observed that among 138 journal figures evaluated from nine leading medical journals, 107 (80%) failed to conform with colour contrast and labelling requirements from the Web Content Accessibility Guidelines (WCAG), indicating that readers with CVD may be unable to perceive displayed information. 215 of 395 (55%) sub-figures within the aforementioned figures were judged to be completely inaccessible to individuals with protan or deutan deficiencies, the commonest CVD subtypes. Despite universal publisher declarations of compliance with WCAG directives, no journals’ figures were fully compliant with these guidelines.

Our findings demonstrate the need for urgent action by authors and publishers to augment the colour contrast of journal figures and add secondary labels to enable their comprehension by audiences with CVD. We believe that these design changes will, too, improve the clarity of figures for general audiences.

## Introduction

Colour vision deficiency (CVD), commonly known as colour blindness, is primarily an X-linked recessive disorder that results from defects in photoreceptor (cone) cells. The condition is estimated to affect more than 300 million individuals worldwide and is more commonly found in males and individuals with Caucasian ancestry. Though heterogeneous in presentation and severity, all types of CVD cause some level of difficulty in distinguishing colour combinations.^1^ Encountering inaccessible learning materials can hinder the educational achievement and progression of individuals with CVD.^2,3^ Moreover, for affected individuals working in medicine and its allied professions, inaccessible academic sources can impede understanding of key research findings, with potential consequences for patients and the wider health system.

The Web Content Accessibility Guidelines (WCAG; https://www.w3.org/WAI/standards-guidelines/wcag/) recommend the use of sufficient colour contrast and non-colour visual indicators (e.g., shape or line type) and labels to distinguish content and facilitate straightforward access to visual information presentations for readers with diverse accessibility needs, including CVD. Though these guidelines are internally recognized, the extent to which major medical journals adhere to these standards in their figures is unknown. Therefore, we conducted a cross-sectional analysis of CVD accessibility in research article figures from nine leading medical journals.

## Methods

We examined figures from the three most recently published research articles as of July 14^th^, 2024 from the top nine journals in the “Medicine” subject area in SCImago’s 2023 journal ranking (https://www.scimagojr.com/journalrank.php?area=2700&year=2023) while excluding articles and journals whose published figures were generally outside the scope of this analysis (such as tables, monochrome figures, or microscope images; **Appendix S1**).

Each figure and graphical abstract in included articles (henceforth known as “plates”), as well as sub-figures within each plate, were evaluated to determine whether they conformed with WCAG 2.1 AA guidelines (or later iterations such as WCAG 2.2), the minimum standard rating to ensure CVD accessibility. Plates were also evaluated to determine whether they would be completely inaccessible to individuals with protan and deutan CVD subtypes (the two commonest CVD subtypes, associated with red and green vision deficiencies respectively).

For analyses of WCAG conformance, we ascertained whether plates and sub-figures contained adequate secondary non-colour indicators of information (i.e., patterns, shapes, and/or labels that themselves met WCAG 2.1 AA in presentation) or employed a colour contrast ratio of 3:1 or greater between all colours intending to be compared. Contrast ratios were determined using the World-Wide-Web-Consortium-recommended Color Contrast Checker (https://www.tpgi.com/color-contrast-checker/). Plates and sub-figures failing to meet either of these conditions were judged to be non-conformant with WCAG guidelines.

For analyses of complete inaccessibility to deutan and protan subtypes, we used the aforementioned Color Contrast Checker’s colour vision simulator tool to judge whether it may be possible for individuals with these common CVD subtypes to access information in figures despite their non-conformance with WCAG guidelines. It should be noted, however, that figures that do not adhere to these guidelines will be difficult and/or impossible to access by individuals with other CVD subtypes and non-CVD visual impairments.

Detailed methods and examples of analyses are provided in **Appendix S2**.

## Results

Characteristics of included journals are described in **Table S2**. Analyses included 138 plates encompassing 395 sub-figures from 27 research articles. In total, 107 (80%) of these plates were partially or wholly non-compliant with WCAG 2.1 AA guidelines on colour contrast or labelling, while 215 (55%) sub-figures within these plates were judged to be completely inaccessible to individuals with common red-green subtypes of CVD. Within all nine journals evaluated, at least 50% of all plates did not comply with WCAG guidelines (**Table 1**).

**Table 1:** CVD accessibility of major medical journals.

| **Journal name** | **Number of issues assessed** | **Number of articles assessed** | **Number of total plates assessed** | **Number of total figures assessed** | **N (%) of plates failing to meet WCAG 2.1 AA guidelines** | **N (%) of total figures completely inaccessible** |
| --- | --- | --- | --- | --- | --- | --- |
| Annals of Oncology | 1 | 3 | 10 | 24 | 10 (100) | 18 (75) |
| CA: A Cancer Journal for Clinicians | 2 | 3 | 34 | 49 | 19 (56) | 13 (27) |
| Cancer Cell | 1 | 3 | 18 | 123 | 18 (100) | 84 (68) |
| Journal of Clinical Oncology | 1 | 3 | 5 | 8 | 4 (80) | 7 (88) |
| Journal of the American College of Cardiology | 1 | 3 | 12 | 27 | 11 (92) | 3 (11) |
| Nature Medicine | 1 | 3 | 26 | 69 | 21 (85) | 28 (41) |
| New England Journal of Medicine | 1 | 3 | 4 | 17 | 3 (75) | 1 (6) |
| Science Immunology | 1 | 3 | 18 | 66 | 18 (100) | 60 (91) |
| The Lancet | 2 | 3 | 6 | 8 | 3 (50) | 1 (13) |
| **Total** | **11** | **27** | **133** | **391** | **107 (80)** | **215 (55)** |

Journals that redrew author-provided figures published fewer inaccessible plates than journals that published unedited figures. Among the latter, journals providing author guidelines related to CVD accessibility (albeit incomplete guidelines such as the avoidance of red and green together) published fewer inaccessible plates (**Table S3**). Though publishers of all analysed journals declared commitments to WCAG 2.0, 2.1, or 2.2 guidelines (**Table S2**), no journals’ figures were fully compliant with these guidelines in terms of CVD accessibility.

## Discussion

Our analyses show that most figures in research articles published by top-ranked medical journals are inaccessible to individuals with CVD. Figures’ inadequate colour contrast and lack of non-colour labelling conflicts with journal publishers’ stated accessibility commitments and may exclude substantial proportions of article audiences.

Advances in assistive technology, including overlays and browser extensions, have allowed a wider variety of digital media to be accessed by some individuals with CVD. However, these tools are unable to render all content accessible and will not work for all individuals with the diagnosis, as shown by user reviews.^4,5^ Moreover, these tools are only likely to be sought out by individuals already diagnosed with CVD, are not applicable to printed articles, and may not change colours in embedded image content. Efforts to ensure the accessibility of journal figures therefore remain paramount.

Simple design changes can help figures fulfil WCAG 2.1 criteria for colour contrast ratios and labelling. We have provided an example of these changes in **Appendix S5**. In recent years, Colour Blind Awareness, a UK-based advocacy organisation, has achieved improvements in the CVD accessibility of selected secondary school assessments (GCSEs and AS and A Levels in England and Wales from Awarding Organisations regulated by Ofqual).^6,7^

We call on scientific publishers and journal editorial boards to lead the way in championing access for readers with CVD, especially given the condition’s substantial prevalence. Journal submission guidelines should espouse the benefits of CVD-friendly content (e.g., a wider potential readership) and provide tutorials to help authors design their figures. At the resubmission stage, online submission systems can offer built-in CVD simulators against which authors can check their article figures.

Journals that redraw author figures in house style prior to publication should revise their visual identities to retain familiar colour palettes, formatting, and fonts while integrating secondary shape- or text-based labels. After publication, journals should offer a standalone forum for readers to raise concerns about the CVD accessibility of figures and correct figures accordingly, a practice that can also be applied to other accessibility issues such as the compatibility of figures with screen-readers. We believe that these changes will “level the playing field” for journal audiences and advance the clarity and impact of published research evidence.

Our analyses are limited by the narrow scope of journals and articles included, though our selection of articles was likely to be random and, therefore, representative of usual journal content. Automated tools for figure accessibility assessment may help future researchers evaluate a greater volume and diversity of journal figures.

## Data Availability

All data analysed in the present work are publicly available.

## Author contributions

YW and KAW conceptualised the study. YW curated publicly available data. KAW performed formal analysis. KAW and YW wrote the original draft and reviewed and edited subsequent drafts.

## Acknowledgements

These analyses were inspired by concerns initially raised by Stelios Boulitsakis Logothetis from the University of Cambridge. The authors would like to thank Professor Trish Greenhalgh from the University of Oxford for granting us permission to use an early version of a published figure to demonstrate the steps required to make figures accessible to individuals with CVD. We also thank Drs Mayank Dalakoti and Holly Pavey from the University of Cambridge for their helpful comments on an earlier version of this article.

