## Supplementary Materials for "Figure accessibility for readers with colour vision deficiency: analysis of leading medical journals"

Kathryn Albany-Ward, BSc (Hons)^1*^

Yaning Wu, MSc^2,3,4*^

1. Colour Blind Awareness, UK
2. British Heart Foundation Cardiovascular Epidemiology Unit, Department of Public Health and Primary Care, University of Cambridge, Cambridge, UK
3. Victor Phillip Dahdaleh Heart and Lung Research Institute, University of Cambridge, Cambridge, UK
4. National Institute for Health and Care Research Blood and Transplant Research Unit in Donor Health and Behaviour, University of Cambridge, Cambridge, UK

*These authors contributed equally.

**Appendix S1: Selection of eligible journals for analysis**

**Table S1: Selection of eligible journals for analysis**

| **Journal** | **Included in analyses?** | **Reason for exclusion** |
| --- | --- | --- |
| Ca-A Cancer Journal for Clinicians | Yes | - |
| Nature Reviews Cancer | No | (Monochrome) tables and illustrations, not graphs, predominate non-text materials |
| Nature Reviews Drug Discovery | No | (Monochrome) tables and illustrations, not graphs, predominate non-text materials |
| Nature Reviews Clinical Oncology | No | (Monochrome) tables and illustrations, not graphs, predominate non-text materials |
| New England Journal of Medicine | Yes | - |
| Nature Medicine | Yes | - |
| Current Protocols in Bioinformatics | No | Screenshots predominate non-text materials (outside scope) |
| MMWR Supplements | No | Colour scheme is monochrome |
| Cancer Cell | Yes | - |
| Nature Reviews Immunology | No | (Monochrome) tables and illustrations, not graphs, predominate non-text materials |
| World Psychiatry | No | Colour scheme is monochrome |
| Annual Review of Immunology | No | (Monochrome) tables and illustrations, not graphs, predominate non-text materials |
| MMWR Recommendations and Reports | No | Colour scheme is monochrome |
| Nature Reviews Genetics | No | (Monochrome) tables and illustrations, not graphs, predominate non-text materials |
| Annals of Oncology | Yes | - |
| Immunity | No | Contains substantial number of micrographs (outside current scope) |
| Morbidity and Mortality Weekly Report | No | Colour scheme is monochrome |
| Nature Reviews Methods Primers | No | (Monochrome) tables and illustrations, not graphs, predominate non-text materials |
| Lancet Oncology | No | Journal in same family with identical colour scheme already included |
| Lancet | Yes | - |
| Nature Cancer | No | Journal in same family with identical colour scheme already included |
| Annual Review of Pathology: Mechanisms of Disease | No | (Monochrome) tables and illustrations, not graphs, predominate non-text materials |
| Nature Immunology | No | Journal in same family with identical colour scheme already included |
| Physiological Reviews | No | Illustrations, not figures, predominate non-text materials |
| Journal of Clinical Oncology | Yes | - |
| MMWR Surveillance Summaries | No | Colour scheme is monochrome |
| Lancet Diabetes and Endocrinology | No | Journal in same family with identical colour scheme already included |
| Nature Reviews Disease Primers | No | (Monochrome) tables and illustrations, not graphs, predominate non-text materials |
| Journal of Hepatology | No | Contains substantial number of micrographs (outside current scope) |
| Nature Reviews Gastroenterology and Hepatology | No | (Monochrome) tables and illustrations, not graphs, predominate non-text materials |
| Nature Reviews Microbiology | No | (Monochrome) tables and illustrations, not graphs, predominate non-text materials |
| Lancet Public Health | No | Journal in same family with identical colour scheme already included |
| Lancet Gastroenterology and Hepatology | No | Journal in same family with identical colour scheme already included |
| Journal of the American College of Cardiology | Yes | - |
| Science immunology | Yes | - |

**Appendix S2: Detailed methods for ascertaining CVD accessibility of journal figures**

1. Each figure was initially assessed to ascertain whether information differentiated by colour was also differentiated by a secondary indicator, such as patterns, shapes, dotted/dashed lines, or additional diagram labels. **Figures S1 and S2** are examples of figures differentiated by text as a secondary indicator. Figures that used secondary indicators which, themselves, employed sufficient contrast with other figure colours in accordance with WCAG 2.1 AA guidelines were judged to conform with those guidelines. Despite the presence of labels as secondary indicators for disease type in **Figure S1**, these labels showed insufficient colour contrast with the background of the figure, meaning that this figure failed to conform with WCAG guidelines. Moreover, given that the lines in Figure S1 intersect, labels at the end of lines alone would have been insufficient to render the figure completely conformant with WCAG guidelines; an additional secondary indicator, such as differing line types, would have been necessary to achieve conformance. On the other hand, **Figure S2** conforms with WCAG guidelines because the labels for educational attainment categories show sufficient colour contrast with the colours of each segment of the donut chart and with the background colour of the figure where the text overlaps with the figure background despite insufficient contrast between the colours of each segment.
2. Where a figure included two or more colours that indicated meaningful information, a W3C-recommended online colour contrast checker (<https://www.tpgi.com/color-contrast-checker/>) was used to ascertain whether all combinations of figure colours that conveyed substantive information achieved a minimum contrast ratio of 3:1. Figures achieving this contrast ratio or higher were judged to conform with WCAG 2.1 AA guidelines. The colours used to denote fruit and vegetable intake categories in **Figure S3** show insufficient contrast with each other, meaning that this figure failed to conform with WCAG guidelines. However, the colours used to denote rural and urban residence in **Figure S4** show sufficient contrast with each other and with the background of the figure, meaning that the figure conforms with WCAG guidelines.
3. For figures that neither provided secondary indicators for information nor achieved a minimum contrast ratio of 3:1 between all colour combinations, an online colour vision simulator (<https://www.tpgi.com/color-contrast-checker/>) was used to determine whether individuals with severe protan and deutan subtypes of CVD would be likely to comprehend the information presented. Figures were judged to be completely inaccessible to individuals with these CVD sub-types if information was impossible to distinguish within the simulator using the available colours, labels, and/or legends. Though **Figure S3A** did not conform with WCAG guidelines due to insufficient colour contrast, we used the colour vision simulator to determine that individuals with these sub-types would be likely to comprehend the information presented (**Figure S3B**).

**Figure S1: Sample journal figure with insufficient colour contrast between line colours and secondary labels with insufficient colour contrast**

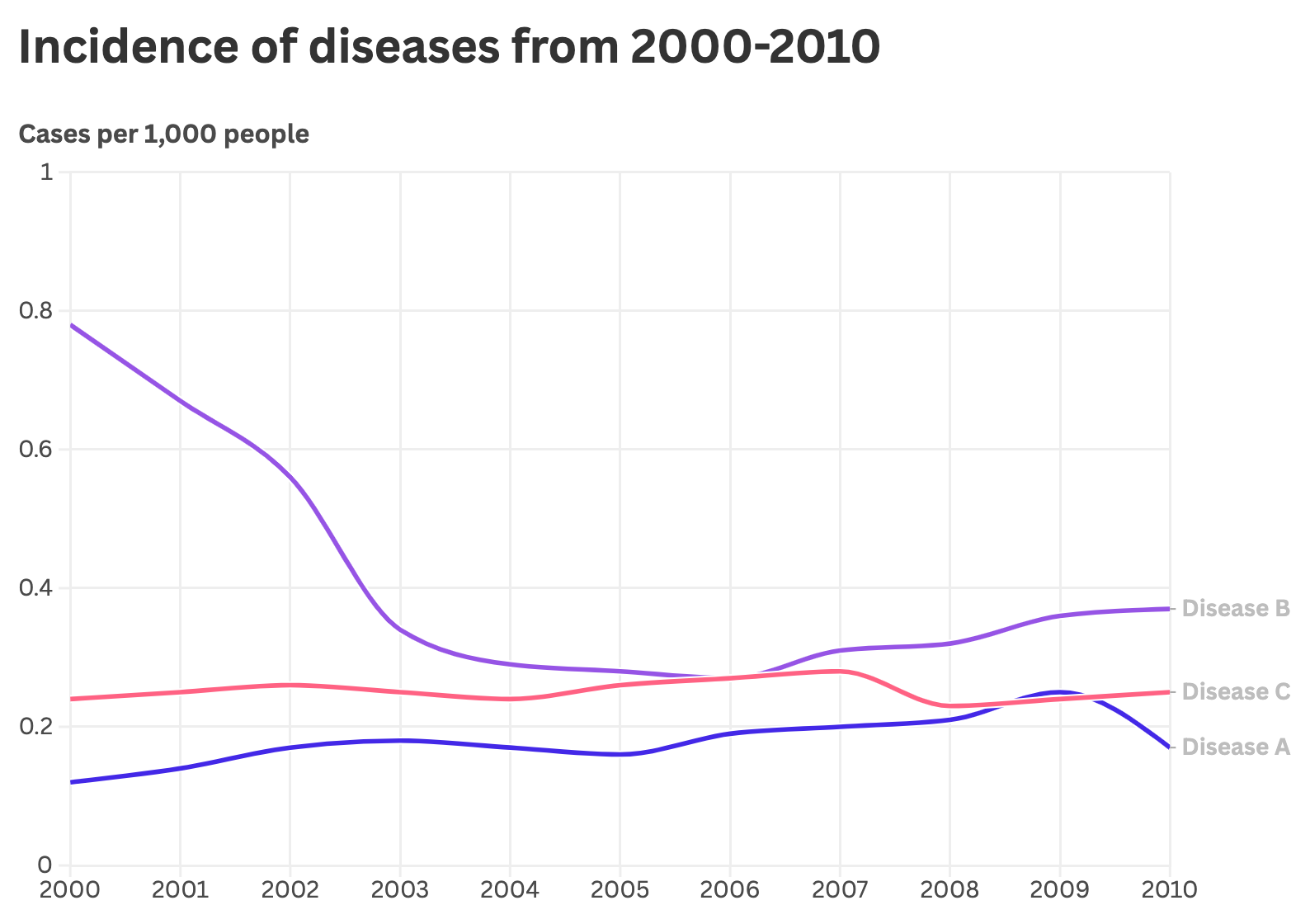

**Figure S2: Sample journal figure with insufficient colour contrast between sector colours but with secondary labels with sufficient colour contrast**

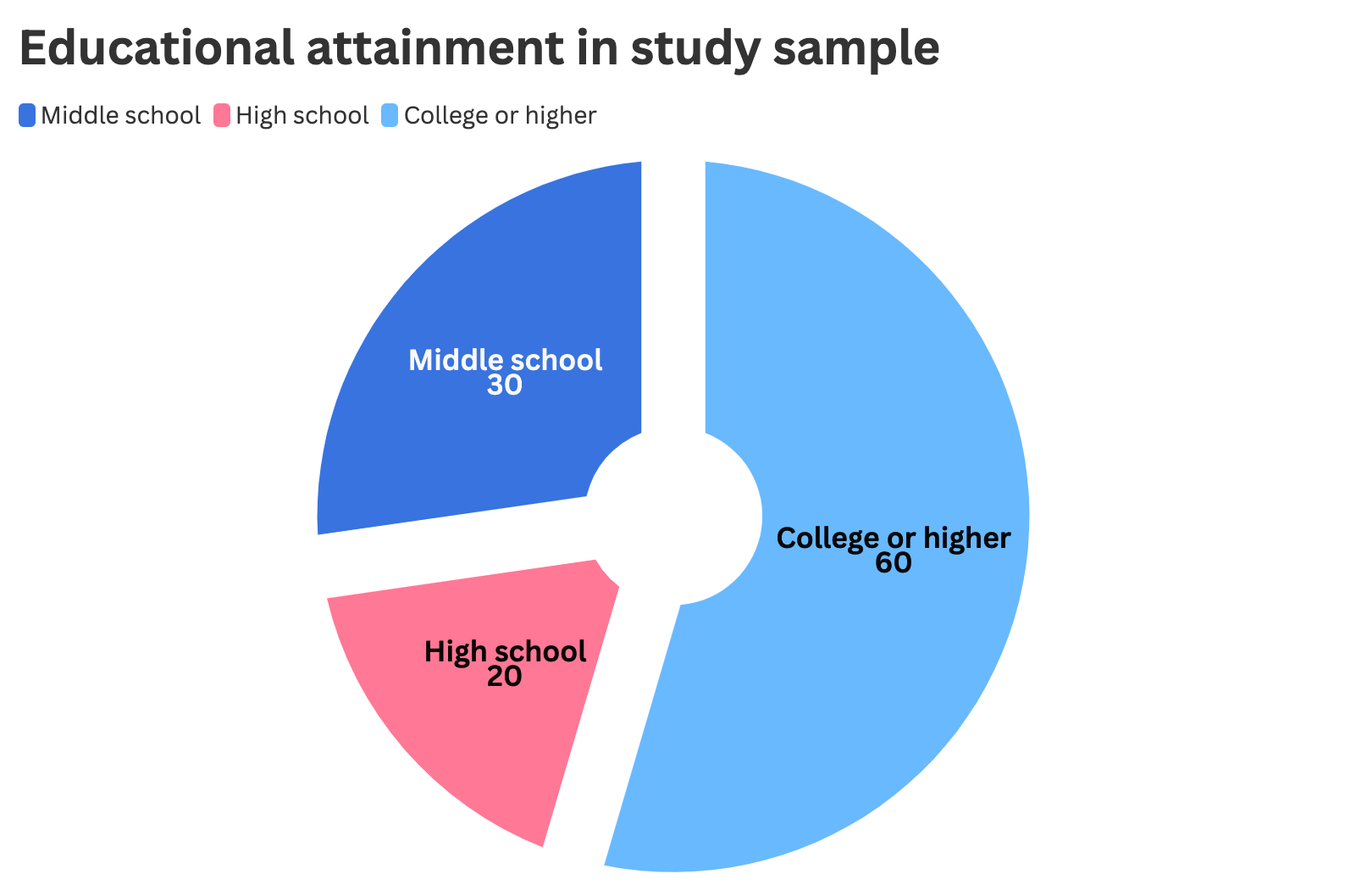

**Figure S3: Sample journal figure without secondary labels and without sufficient colour contrast, though with sufficient colour contrast to be potentially accessible to individuals with protan/deutan CVD subtypes**

**A** Original figure
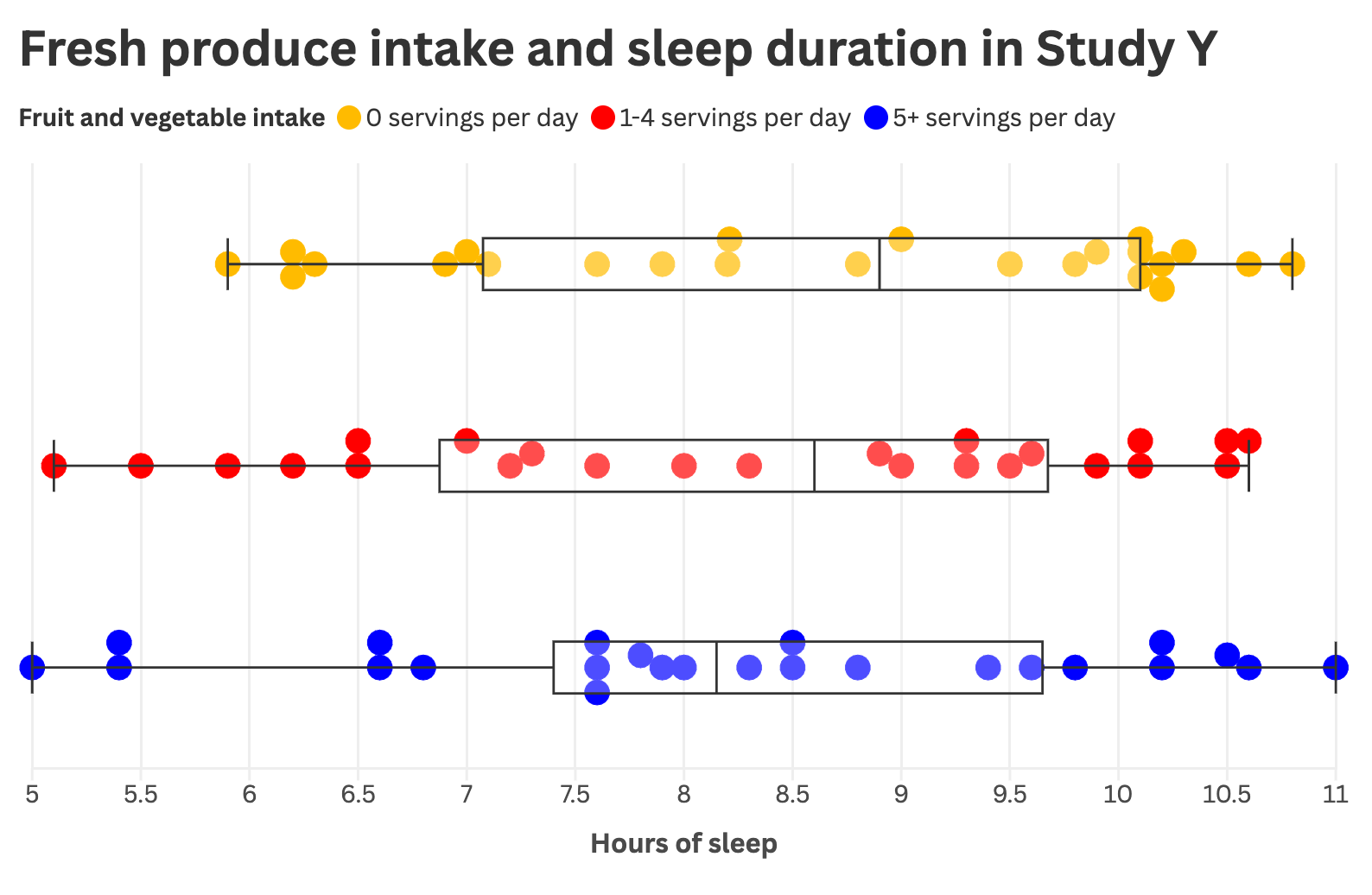

**B** Figure inputted into CVD simulation software for deutan CVD subtypes

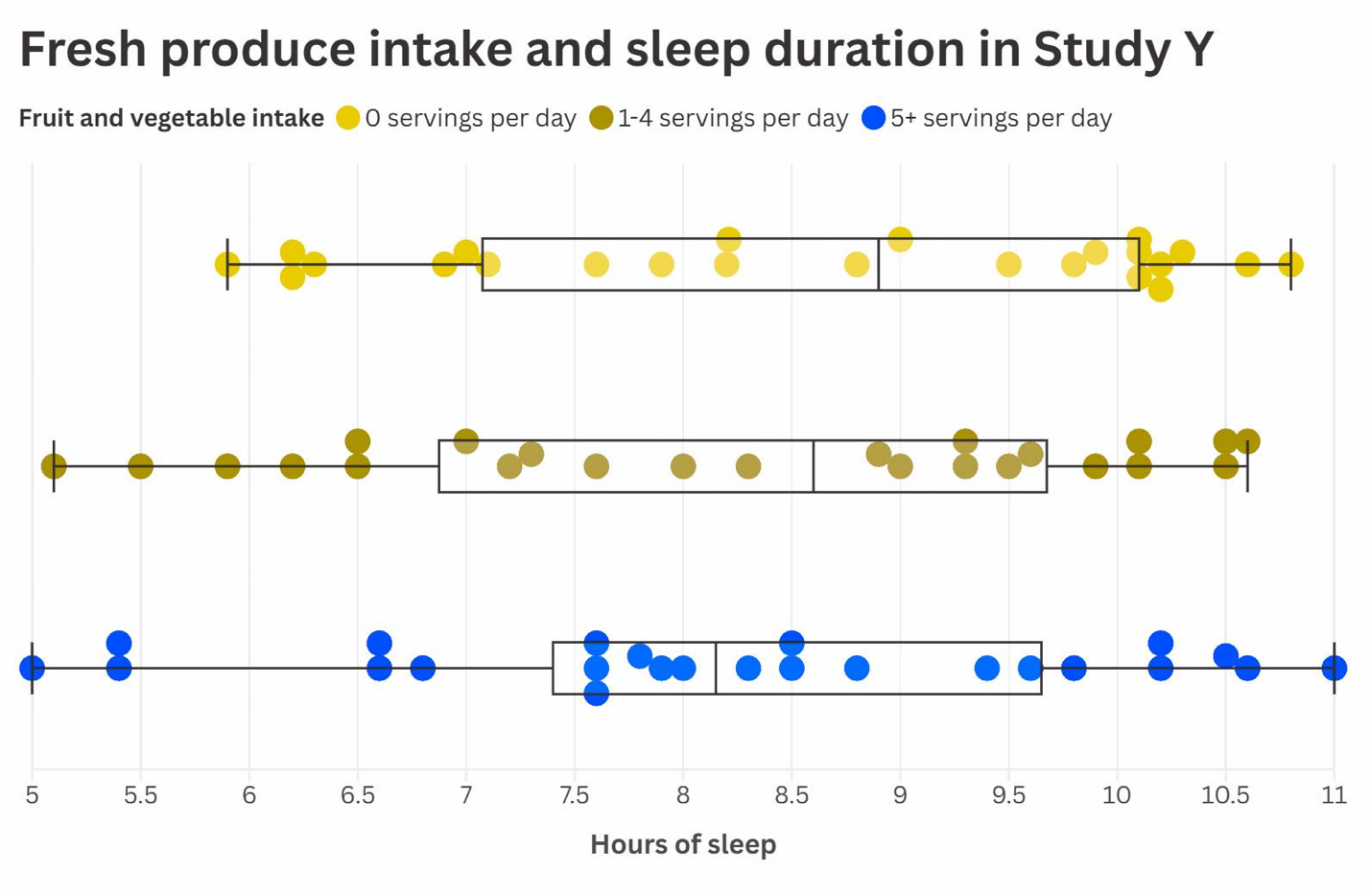

**Figure S4: Sample journal figure without secondary labels but with sufficient colour contrast**

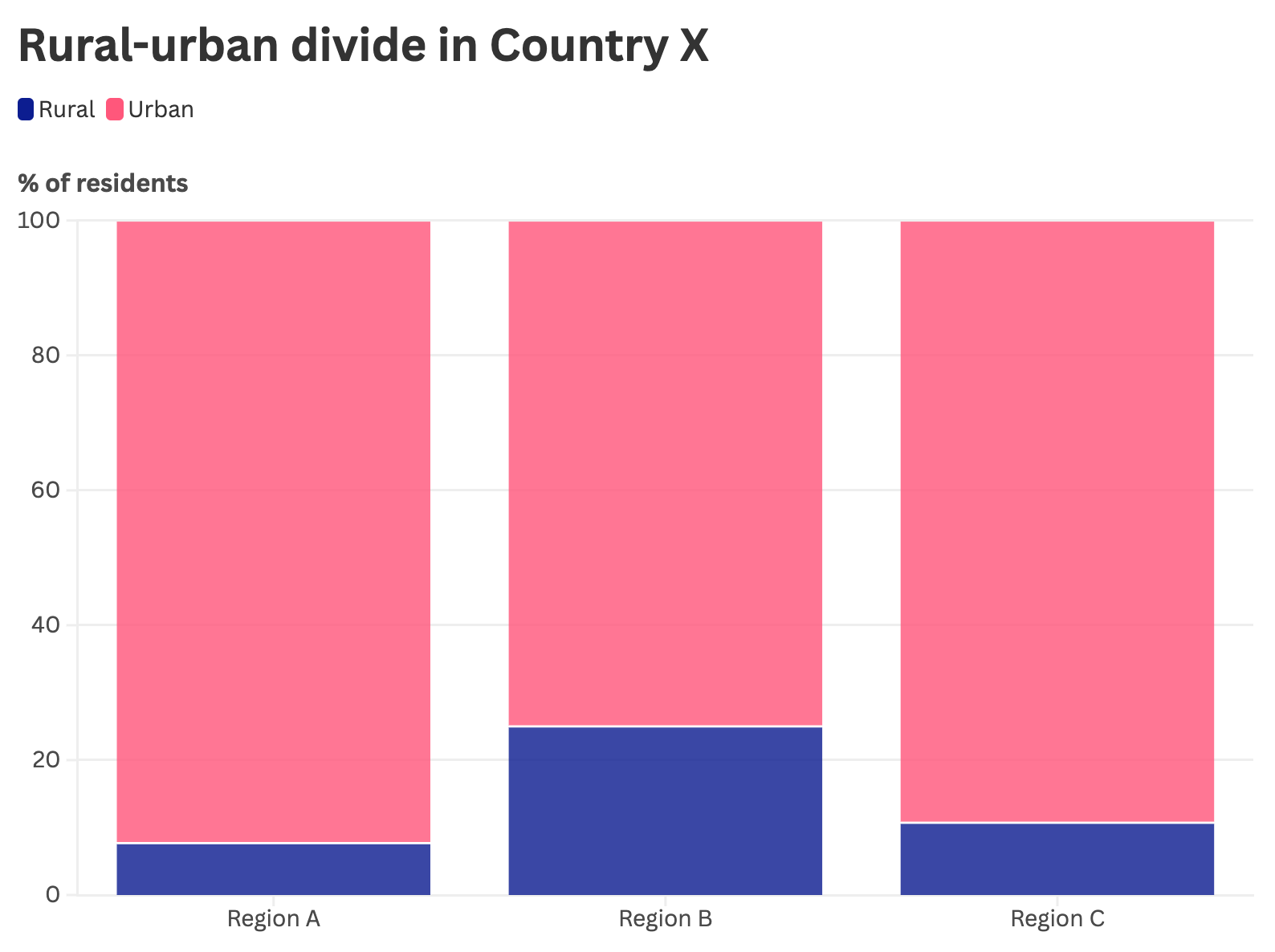

**Appendix S3: Journal descriptions**

**Table S2: General and accessibility-related characteristics of journals included in analyses**

| **Journal name** | **Publisher** | **Accessibility statement link** | **WCAG in accessibility statement?** | **CVD guidelines?** | **Editors redraw?** |
| --- | --- | --- | --- | --- | --- |
| Annals of Oncology | Elsevier | <https://www.elsevier.com/en-gb/about/accessibility> | Y; 2.1 AA | N | N |
| CA: A Cancer Journal for Clinicians | John Wiley & Sons | <https://onlinelibrary.wiley.com/accessibility> | Y; 2.2 A/AA | Y (?) | N |
| Cancer Cell | Cell Press | <https://www.cell.com/accessibility> | Y; 2.0 | Y | N |
| Journal of Clinical Oncology | Lippincott Williams & Wilkins | <https://download.lww.com/vpat/wk-thepoint-accessibility.pdf> | Y; 2.1 | N | N |
| Journal of the American College of Cardiology | Elsevier | <https://www.elsevier.com/en-gb/about/accessibility> | Y; 2.1 | N | Y |
| Nature Medicine | Springer Nature | <https://www.nature.com/info/accessibility-statement> | Y; 2.1 | N | Y |
| New England Journal of Medicine | Massachusetts Medical Society | <https://www.nejm.org/about-nejm/frequently-asked-questions?#Accessibility> | Y; 2.0 | N | Y |
| Science Immunology | American Association for the Advancement of Science | <https://www.science.org/content/page/accessibility> | Y; 2.1 | Y | N |
| The Lancet | Elsevier | <https://www.elsevier.com/en-gb/about/accessibility> | Y; 2.1 | N | Y |

**Appendix S4: Supplementary results**

**Table S3: Subgroup analyses of CVD accessibility by journal characteristics**

| **Journal type** | **Number of journals** | **N (%) of plates failing to meet WCAG 2.2AA guidelines** | **N (%) of total figures completely inaccessible** |
| --- | --- | --- | --- |
| Figures re-drawn | 4 | 38 (79) | 33 (27) |
| Figures not re-drawn | 6 | 69 (81) | 182 (67) |
| Any CVD guidelines | 3 | 55 (78) | 157 (66) |
| No CVD guidelines (all) | 7 | 52 (82) | 58 (38) |
| No CVD guidelines (figures not re-drawn) | 2 | 14 (93) | 25 (78) |

**Appendix S5: Methods for improving CVD accessibility of an example figure**

We illustrate one potential method for improving the accessibility of journal figures to readers with CVD using an earlier, unpublished version of a journal table from Jones and colleagues’ 2020 publication**^1^** (**Table S4**), which shows levels of COVID-19 transmission risk associated with various activities and face covering use during the pandemic. Permission for our reproduction of the table was kindly granted by one of the publication's senior authors, Professor Trish Greenhalgh. These accessibility enhancements were originally devised and shared by the non-profit organisation Colour Blind Awareness.

**Table S4**: Original unpublished version of journal figure from Jones et al^1^

| **Wearing face coverings, contact for a short time** | | | | | | | |
| --- | --- | --- | --- | --- | --- | --- | --- |
|  | Low occupancy | | |  | High occupancy | | |
|  | Outdoors, well ventilated | Indoors, well ventilated | Poorly ventilated |  | Outdoors, well ventilated | Indoors, well ventilated | Poorly ventilated |
| Silent |  |  |  |  |  |  |  |
| Speaking |  |  |  |  |  |  |  |
| Shouting, singing |  |  |  |  |  |  |  |

| **Wearing face coverings, contact for a prolonged time** | | | | | | | |
| --- | --- | --- | --- | --- | --- | --- | --- |
|  | Low occupancy | | |  | High occupancy | | |
|  | Outdoors, well ventilated | Indoors, well ventilated | Poorly ventilated |  | Outdoors, well ventilated | Indoors, well ventilated | Poorly ventilated |
| Silent |  |  |  |  |  |  |  |
| Speaking |  |  |  |  |  |  |  |
| Shouting, singing |  |  |  |  |  |  |  |

| **No face coverings, contact for a short time** | | | | | | | |
| --- | --- | --- | --- | --- | --- | --- | --- |
|  | Low occupancy | | |  | High occupancy | | |
|  | Outdoors, well ventilated | Indoors, well ventilated | Poorly ventilated |  | Outdoors, well ventilated | Indoors, well ventilated | Poorly ventilated |
| Silent |  |  |  |  |  |  |  |
| Speaking |  |  |  |  |  |  |  |
| Shouting, singing |  |  |  |  |  |  |  |

| **No face coverings, contact for a prolonged time** | | | | | | | |
| --- | --- | --- | --- | --- | --- | --- | --- |
|  | Low occupancy | | |  | High occupancy | | |
|  | Outdoors, well ventilated | Indoors, well ventilated | Poorly ventilated |  | Outdoors, well ventilated | Indoors, well ventilated | Poorly ventilated |
| Silent |  |  |  |  |  |  |  |
| Speaking |  |  |  |  |  |  |  |
| Shouting, singing |  |  |  |  |  |  |  |

| Risk of transmission | low |  | medium |  | high |
| --- | --- | --- | --- | --- | --- |

**Table S5** shows a simulation of how **Table S4** would likely appear to individuals with protanopia – the risk categories differentiated in green, yellow, and red (“low”, “medium”, and “high”) are generally indistinguishable.

**Table S5: Simulation of view of Table S4 for individuals with severe protan CVD subtype**

| **Wearing face coverings, contact for a short time** | | | | | | | |
| --- | --- | --- | --- | --- | --- | --- | --- |
|  | Low occupancy | | |  | High occupancy | | |
|  | Outdoors, well ventilated | Indoors, well ventilated | Poorly ventilated |  | Outdoors, well ventilated | Indoors, well ventilated | Poorly ventilated |
| Silent |  |  |  |  |  |  |  |
| Speaking |  |  |  |  |  |  |  |
| Shouting, singing |  |  |  |  |  |  |  |

| **Wearing face coverings, contact for a prolonged time** | | | | | | | |
| --- | --- | --- | --- | --- | --- | --- | --- |
|  | Low occupancy | | |  | High occupancy | | |
|  | Outdoors, well ventilated | Indoors, well ventilated | Poorly ventilated |  | Outdoors, well ventilated | Indoors, well ventilated | Poorly ventilated |
| Silent |  |  |  |  |  |  |  |
| Speaking |  |  |  |  |  |  |  |
| Shouting, singing |  |  |  |  |  |  |  |

| **No face coverings, contact for a short time** | | | | | | | |
| --- | --- | --- | --- | --- | --- | --- | --- |
|  | Low occupancy | | |  | High occupancy | | |
|  | Outdoors, well ventilated | Indoors, well ventilated | Poorly ventilated |  | Outdoors, well ventilated | Indoors, well ventilated | Poorly ventilated |
| Silent |  |  |  |  |  |  |  |
| Speaking |  |  |  |  |  |  |  |
| Shouting, singing |  |  |  |  |  |  |  |

| **No face coverings, contact for a prolonged time** | | | | | | | |
| --- | --- | --- | --- | --- | --- | --- | --- |
|  | Low occupancy | | |  | High occupancy | | |
|  | Outdoors, well ventilated | Indoors, well ventilated | Poorly ventilated |  | Outdoors, well ventilated | Indoors, well ventilated | Poorly ventilated |
| Silent |  |  |  |  |  |  |  |
| Speaking |  |  |  |  |  |  |  |
| Shouting, singing |  |  |  |  |  |  |  |

| Risk of transmission | low |  | medium |  | high |
| --- | --- | --- | --- | --- | --- |

In rendering this table more accessible to individuals with CVD, we sought to preserve the “traffic light” risk categorisation system used in the original table given general audiences' familiarity with these colours and their connotation.

Because it was not possible to identify three distinct colours that fulfilled the WCAG-2.1-required 3:1 contrast ratio with each other and with both white and black, we used shades of green, yellow, and red that fulfilled a 3:1 contrast ratio with either white or black (but not necessarily with each other), added secondary labels of “L”, “M”, and “H” to denote risk categories in each cell of the table, and retained the black outlining used in the original table to preserve sufficient contrast between all table elements. The resulting table, **Table S6**, is accessible to individuals with all types and severities of CVD and retains the “traffic light” coding commonly understood by general audiences.

**Table S6: CVD-accessible version of Table S4**

| **Wearing face coverings, contact for a short time** | | | | | | | |
| --- | --- | --- | --- | --- | --- | --- | --- |
|  | Low occupancy | | |  | High occupancy | | |
|  | Outdoors, well ventilated | Indoors, well ventilated | Poorly ventilated |  | Outdoors, well ventilated | Indoors, well ventilated | Poorly ventilated |
| Silent | L | L | **L** |  | L | L | M |
| Speaking | L | L | L |  | L | L | M |
| Shouting, singing | L | L | M |  | M | M | H |

| **Wearing face coverings, contact for a prolonged time** | | | | | | | |
| --- | --- | --- | --- | --- | --- | --- | --- |
|  | Low occupancy | | |  | High occupancy | | |
|  | Outdoors, well ventilated | Indoors, well ventilated | Poorly ventilated |  | Outdoors, well ventilated | Indoors, well ventilated | Poorly ventilated |
| Silent | L | L | M |  | L | M | H |
| Speaking | L | L | M |  | M | M | H |
| Shouting, singing | L | M | H |  | M | H | H |

| **No face coverings, contact for a short time** | | | | | | | |
| --- | --- | --- | --- | --- | --- | --- | --- |
|  | Low occupancy | | |  | High occupancy | | |
|  | Outdoors, well ventilated | Indoors, well ventilated | Poorly ventilated |  | Outdoors, well ventilated | Indoors, well ventilated | Poorly ventilated |
| Silent | L | L | M |  | M | M | H |
| Speaking | L | M | M |  | M | H | H |
| Shouting, singing | M | M | H |  | H | H | H |

| **No face coverings, contact for a prolonged time** | | | | | | | |
| --- | --- | --- | --- | --- | --- | --- | --- |
|  | Low occupancy | | |  | High occupancy | | |
|  | Outdoors, well ventilated | Indoors, well ventilated | Poorly ventilated |  | Outdoors, well ventilated | Indoors, well ventilated | Poorly ventilated |
| Silent | L | M | H |  | M | H | H |
| Speaking | M | M | H |  | H | H | H |
| Shouting, singing | M | H | H |  | H | H | H |

| Risk of transmission | low | L | medium | M | high | H |
| --- | --- | --- | --- | --- | --- | --- |
